# Facilitators and Barriers for the use of Direct Anticoagulant: A qualitative study

**DOI:** 10.1101/2025.10.01.25336915

**Authors:** Puspanjali Adhikari, Yunika Acharya, Sneha Shrestha, Manisha Makaju, Abiodun Adeoye, Di Cesare Mariachiara, Biraj Man Karmacharya

**Author notes:** Corresponding author: Di Cesare Mariachiara.

## Abstract

**Introduction:** Atrial fibrillation (AF) is the most common arrhythmia globally and a leading cause of stroke, heart failure, and cardiovascular disease. In low- and middle-income countries (LMICs), including Nepal, managing AF is challenging due to limited access to timely diagnosis and traditional treatments like Vitamin K Antagonists (VKAs). Non-Vitamin K antagonist oral anticoagulants (NOACs) have improved stroke prevention in AF patients, but affordability and accessibility remain significant barriers. This study explores barriers and facilitators to NOAC use for stroke prevention among AF patients in Nepal.

**Materials and Methods:** This qualitative study involved in-depth interviews with 57 stakeholders in cardiovascular disease programs, policies, and management in Bagmati province, Nepal. Interviews were digitally recorded, transcribed verbatim, and analyzed using NVivo software.

**Results:** Barriers included low awareness among healthcare providers regarding NOACs rural health workers and patients, lack of essential equipment, insufficient human resources, absence of dedicated policies, and limited specialized training. The cost and unavailability of advanced medications like NOACs further hindered access. Challenges in Prothrombin time Internalized Normalized Ration(INR) monitoring for Warfarin users also persisted. Key facilitators were increased awareness among healthcare workers about AF at macro urban levels and the implementation of the Package of Essential Noncommunicable (PEN) disease interventions protocol for managing cardiovascular diseases.

**Conclusion:** This study identifies critical gaps in AF management in Nepal, particularly in rural settings, where limited infrastructure, inadequate training, and low public awareness hinder effective diagnosis and treatment. The high cost and restricted access to NOACs, coupled with challenges in using Warfarin, exacerbate these issues. Addressing these gaps through national guidelines, improved infrastructure, training programs, and cost-effective policies is essential. Embedding an educational program in a broader quality improvement initiative can enhance healthcare workers’ knowledge and ensure equitable and effective AF management across Nepal.

## Introduction

Atrial fibrillation is the most common form of arrhythmia worldwide and is one of the leading causes of stroke, heart failure, sudden deaths, and cardiovascular disease.(1) This results in high healthcare costs and a significant public health burden with over 37.5 million people affected by AF.(2) Between 1990 and 2017, there has been rapid increase in the number of people with atrial fibrillation worldwide, rising from 19.1 million to 37.6 million, and is expected to increase further by > 60% by 2050. An estimated 17.9 million people in low- and middle-income countries (LMICs) will be living with Non Valvular Atrial Fibrillation (NVAF) by 2020. An increasing trend of AF in low resource settings countries with limited access to preventive measures, timely diagnosis, and challenges of regular monitoring with Vitamin K Antagonist (VKSs) oral anticoagulants leads to significant health and economic burden(3)

A systematic review including over 28,000 participants, showed how effective treatment with Vitamin K Antagonist (VKAs) oral anticoagulant medications reduced the risk of stroke in people with AF by two-thirds.(4) However, patients treated with VKAs must be carefully monitored for INR 2.0-3.0 to effectively reduce the risk of embolic stroke while minimizing bleeding risks.(5) Furthermore, factors like alcohol consumption, intake of vitamin K-rich foods and illness interferes with the patients’ ability to stay within the therapeutic range.(6) The regular monitoring required for VKA treatment is particularly challenging for patients and health systems, especially in remote and resource-limited settings where health systems are often poorly equipped to care for people affected by AF. (7,8)

In the past decade there has been a paradigm shift in the treatment of AF with the introduction of Non-Vitamin K antagonist oral anticoagulants (NOACs), a class of medicines which includes Dabigatran, Apixaban, Edoxaban and Rivaroxaban, has improved the safety and efficacy of NVAF (Non Valvular Atrial Fibrillation) treatment for stroke prevention, with significant reductions in stroke, intracranial hemorrhage, and mortality.(9) As opposed to VKAs, NOACs do not require routine INR) testing and have far fewer drug-drug and drug-food interactions.(9)

Considering the advantage of NOACs over VKAs major clinical practice guidelines worldwide recommend NOACs over VKAs for initial treatment of NVAF for stroke prevention.(10) There has been a rapid uptake of NOACs amongst high income countries, with widespread accessibility across all socio-demographic groups.(11) However, the picture is different in low and middle-income countries, where access to NOACs is governed by an inability to afford to pay for treatment.(12)(8) The current availability of NOAC in Nepal along with the lack of clinical experience among physicians, higher cost, unavailability of antidotes, and contraindications in patients with severe kidney or liver disease might be the reason for the lower use of NOACs (e.g. Dabigatran) (3.78%) than VKAs (Warfarin), which has been traditionally used in monotherapy as well as in combination therapy.(13)

After a preliminary failed attempt to include NOACs into the WHO EML undertaken in 2014, based on concern that the application’s supporting data came almost exclusively from RCTs, and underrepresented trial populations there has been a substantial increase in real-world data published with large-scale registry studies, databases of insurance claims, and systematic reviews and meta-analysis of NOACs in special-risk populations (e.g. elderly, renal impairment, and high HAS-BLED score). The introduction of NOAC-specific antidotes and increasing data on costs and cost-effectiveness led to the successful application for the inclusion of NOACs (Dabigatran as representative of the pharmacological class) into the 21st WHO EML, led by the World Heart Federation Emerging Leaders GOALPoST (Improving Global Access to Oral Anticoagulants to Prevent <u>S</u>troke in ATrial fibrillation) team(14)

There is a disparity in NOAC use, with rapid uptake in high-income countries across all socio-demographic groups, while low- and middle-income countries face limited access due to affordability issues. This highlights the need for a health system assessment to improve stroke prevention in AF patients and identify areas to enhance NOAC access and care. (15)

RAPs (Retention and Adherence Programs) have been extensively used for the assessment of health services for communicable diseases like HIV-AIDS(16) and increasingly for non-communicable diseases like diabetes. RAPs allow for generating comprehensive assessments in specific areas of public health in a relatively short time and with fairly low costs, being a valuable tool in resource scarce settings, and are powerful instruments for quantifying and identifying the gaps between clinical evidence and practice. RAPs methodological principles include the use of multiple methods (qualitative and quantitative), multiple data sources (primary and secondary data), and a triangulation between these focusing, at the same time, on the social, cultural and economic contexts in which populations, individuals, and their behaviors are situated.

In particular RAPs allow overcoming one of the main challenges in implementation research - the engagement with the stakeholders (individuals, healthcare givers, and policy makers). This will help to determine the five dimensions of access to medications: availability, affordability, accessibility, acceptability and quality of medicines. Therefore, the objective of this study was to develop the Rapid Assessment Protocol for Direct Oral Anticoagulants access and use (RAP-NOACs) so that it can be applied in Nepal to determine the barriers and facilitators to use of NOACS in stroke prevention in patients with AF.

## Methods

### Study design

This was an exploratory qualitative study conducted among key stakeholders involved in cardiovascular disease programs, policies, and management. We focused on In Depth Interviews (IDIs). The IDIs were categorized across macro, meso, and micro levels to capture perspectives from central, urban, and rural healthcare settings in Bagmati Province, Nepal.

### Study site

The study was conducted in Bagmati Province of Nepal, incorporating central, urban, and rural healthcare settings. The IDIs were systematically categorized across macro, meso, and micro levels to capture multi-tiered perspectives. Macro level: Included national-level institutions such as the Division of Curative Health Services within the Ministry of Health, Cardiac Society of Nepal, Nepal Heart Foundation, Society of Internal Medicine Doctors of Nepal, Neurosurgical Society of Nepal, Nepal Pharmacists’ Association, Nepal Association of Medicine Suppliers, Department of Drug Administration (DDA), and central hospitals. Meso level: Included provincial stakeholders such as the Provincial Ministry of Health, Province Medical Store, Province Hospital, and medical and pharmacology professionals involved in academia and research. Micro level: Included patients, health care workers, and community or traditional healers from both urban and rural communities.

### Sampling procedure

Participants were selected using a purposive sampling approach. A list of potential stakeholders was developed through the extended professional networks of the primary author and co-authors. A list of potential participants was mapped from the extended professional networks of the primary author and co-authors. All the participants were contacted through multiple channels including phone calls, emails and in person reach, depending upon their context and availability. Written informed consent was obtained prior to each interview.

### Developing interview guide

The semi-structured questionnaire guide was developed for the RAP-NOACs was adapted taking into reference The Rapid Assessment Protocol for Insulin Access, which allowed identification of specific paths necessary for the development of the questionnaires to be complemented by the evidence gathered.(17) The questionnaires were further reviewed by national and international experts in the field. The questionnaire was translated and back translated to ensure accuracy and clear understanding. The preliminary RAP-based tool was piloted among 11 participants representing various stakeholders in three hospitals in Nepal between January to February 2020. Based on the findings of the pilot study the initial semi-structured guide was revised, themes and subthemes (Table 2) were developed and a codebook was generated.

Four themes were developed:

1. General knowledge and awareness of AF management
2. Health infrastructure and training to manage AF
3. Policy environment for cost effective management of AF with NOACs
4. Barriers and facilitators to use of NOACs

Questions were open-ended with probing questions prompted by participants’ responses. Participants were given the opportunity to discuss points identified but not outlined in the guide. Individual interviews were conducted by the trained research assistants and project lead. Field notes were also made on interviewees’ emotions, facial expressions and speaking tones during the interviews.

### Data management and analysis

The interviews were digitally recorded, transcribed verbatim and anonymized. The text generated from the transcript were divided into meaningful units, such as phrases and quotes, and the meaningful units were condensed, abstracted and labeled with codes independently by two of the investigators. The various codes were compared based on differences and similarities and sorted into categories. The categories were discussed, codes were grouped and organized into an analytic framework in the form of final themes and sub-themes of two investigators. Transcripts from the interviews were uploaded and analyzed in NVivo (QSR International Pty Ltd., Version 12, 2019, Victoria, Australia) to facilitate the identification and refinement of patterns and themes.

### Ethical considerations

Ethical clearance from the Ethical Review Board of the Nepal Health Research Council (NHRC). Participants were informed that confidentiality would be maintained during the research. Written informed consent was obtained prior to each interview.

## Results

### Theme 1: General Knowledge and Awareness of AF Management

The medical practitioners in the urban advanced setting, health care practitioners from different professional societies and health professionals at policy level knew AF is a condition that causes imbalance in the rhythm of heart, however the health practitioners in the rural settings, pharmacists and patients could only relate AF as a cardiac disease. The rural health settings have provision of managing simple cases like management of AF has been in line of controlling the rhythm primarily and mostly prescribing VKAs. The health practitioners in rural settings were unaware of NOACs as compared to Warfarin, they tended to focus more on treating hypertension than addressing specific cases of AF. One possible reason for this could be the unavailability of basic ECG machines in rural settings, making it difficult to diagnose AF. There are no separate protocols or guidelines for the management of AF. The AF cases are managed based on the individual hospital protocol or international AF management guideline in the urban settings. Majority of the patients know they have heart disease but are unaware that they have AF and would buy medicine based on what is written on the prescription and do not know what the medicine is. **(Table 3)**

**Table 1:** Number of IDIs.

|  | National level | Urban | Rural |
| --- | --- | --- | --- |
| **Macro level** |  |  |  |
| Ministry | 1 |  |  |
| Professional societies | 2 |  |  |
| Central Medical store |  | 4 |  |
| DDA | 2 |  |  |
| Central Hospitals |  | 2 |  |
| **Meso level** |  |  |  |
| Province ministry of health | 1 |  |  |
| Province medical store | 2 |  |  |
| Province Hospital | 3 |  |  |
| Academia/Research |  | 11 |  |
| Pharmacy | 1 | 2 |  |
| **Micro level** |  |  |  |
| Patient | 5 | 5 |  |
| Health Care Worker<br>(Community + Traditional) |  | 4 | 12 |
| **Total** | **17** | **28** | **12** |

**Table 3:**
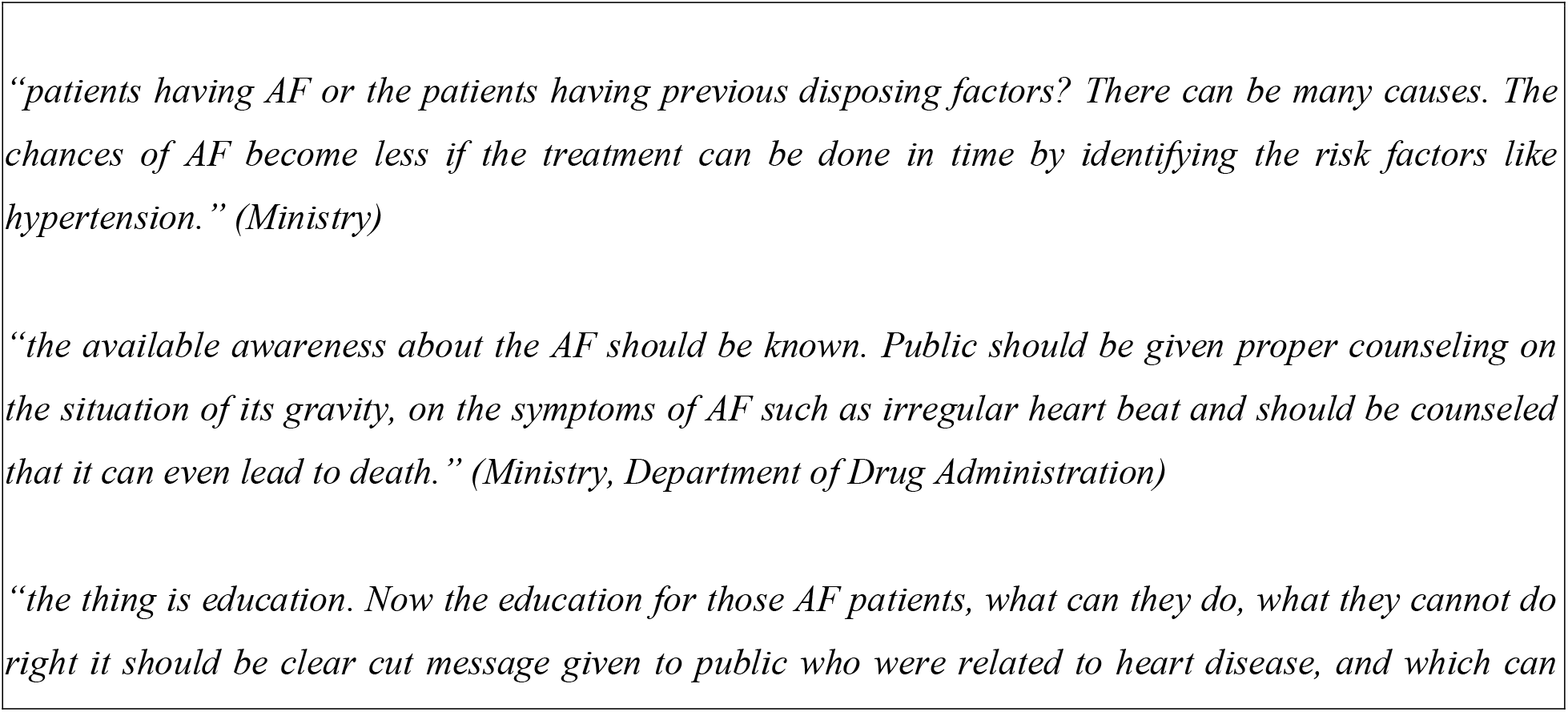
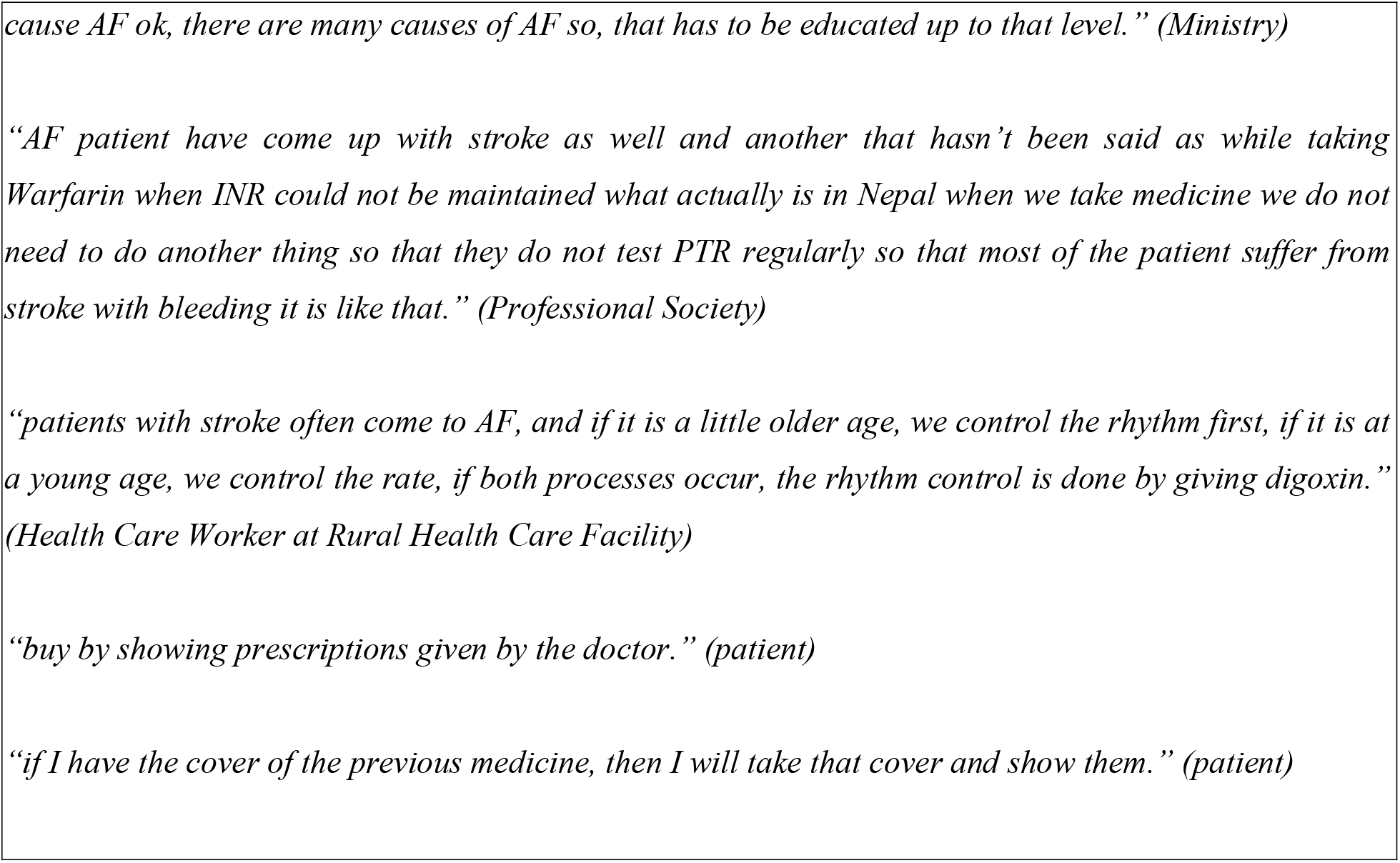
Example Quotes Describing General Knowledge and Awareness of AF Management.

One of the participants (macro level) said *“from a register drugstore where medicine for AF is available, the doctor or practitioner has to explain everything and explain it to the patient about pros and cons of medication. If that is the case then the other one needs to be accompanied by the laboratory facility and the other is the up to date laboratory tests, that feature also had to be fulfilled, and he had to be on his regular medication, becoming aware of the consequences that medicine had to do with everything the drug demanded*.*” (Ministry)* which highlights the lack of proper infrastructure like a lab for INR monitoring and unawareness of the patient about their heart condition and medication usage. It also emphasizes how a healthcare professional should be aware about AF and drugs for its management for proper counseling of the patients.

The patients are not aware and do not understand the importance of INR monitoring according to one of the participants (meso level) *“umm!!what I think is, there is little knowledge about atrial fibrillation in the public level as other diseases. Even When we say to our parents about AF they ask what is it? when they have atrial fibrillation and it is challenging to tell them what is this and how to manage and you have to take care of INR monitoring. It is very difficult for them to understand and even if we explain these in layman’s terms. I think there is a challenge in monitoring the INR regularly”. (Health Care Worker at Rural Health Care Facility)*

### Theme 2: Health Infrastructure, Guidelines and Training to Manage AF

The infrastructure in the rural government setting is not adequate to manage AF where health care providers can manage raised BP with limitation to diagnose AF because they lack the basic equipment like ECG. There is lack of medical doctors at Primary Health Care Level, which results in these centers lacking lab or equipment to diagnose AF. The doubtful cases of AF get refereed to tertiary or cardiac specialized centers where there is no provision of free treatment for AF. The health care practitioners in both rural and urban settings do not have a separate training for management of AF and refer to different international guidelines. **(Table 4)**

**Table 4:**
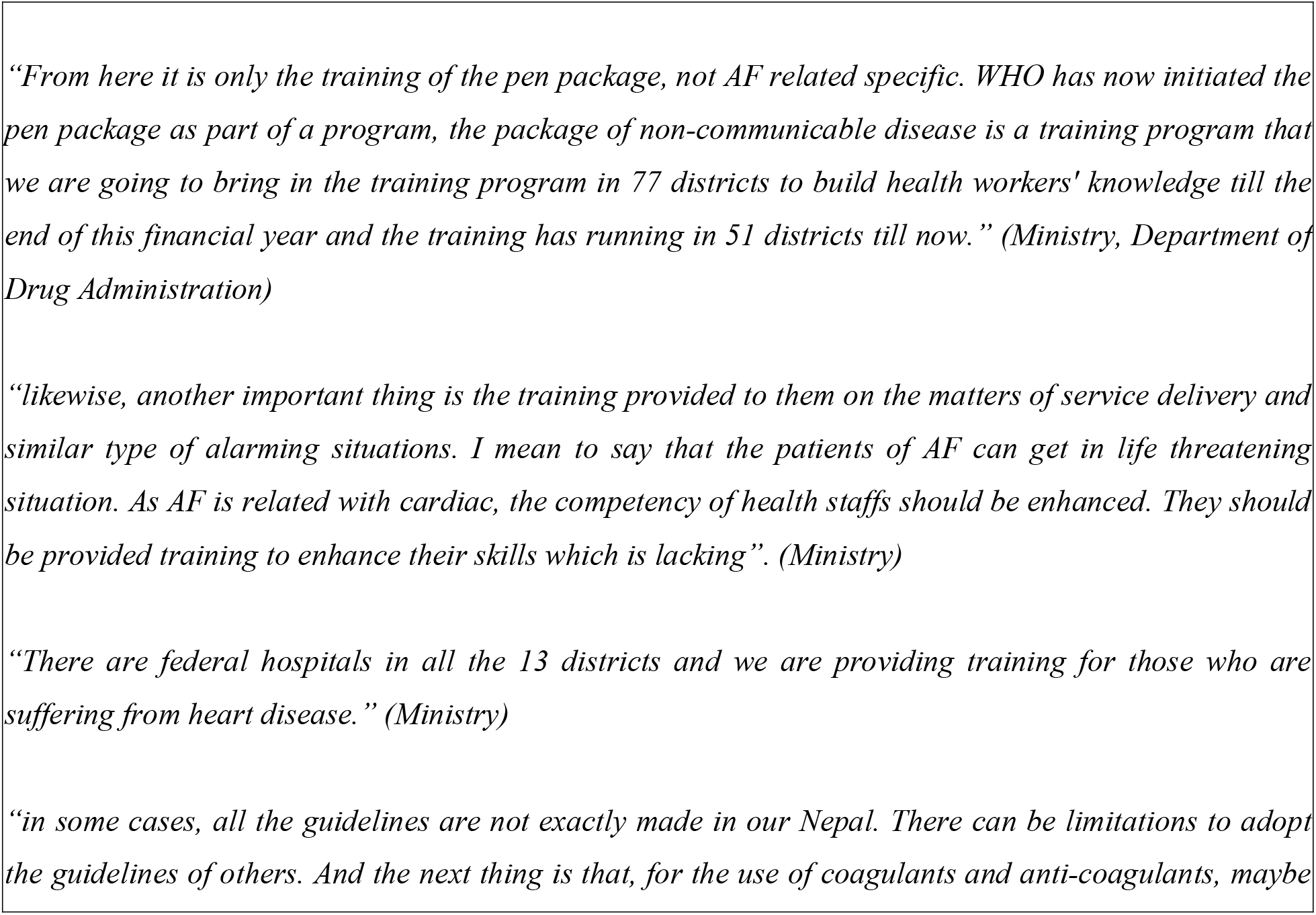
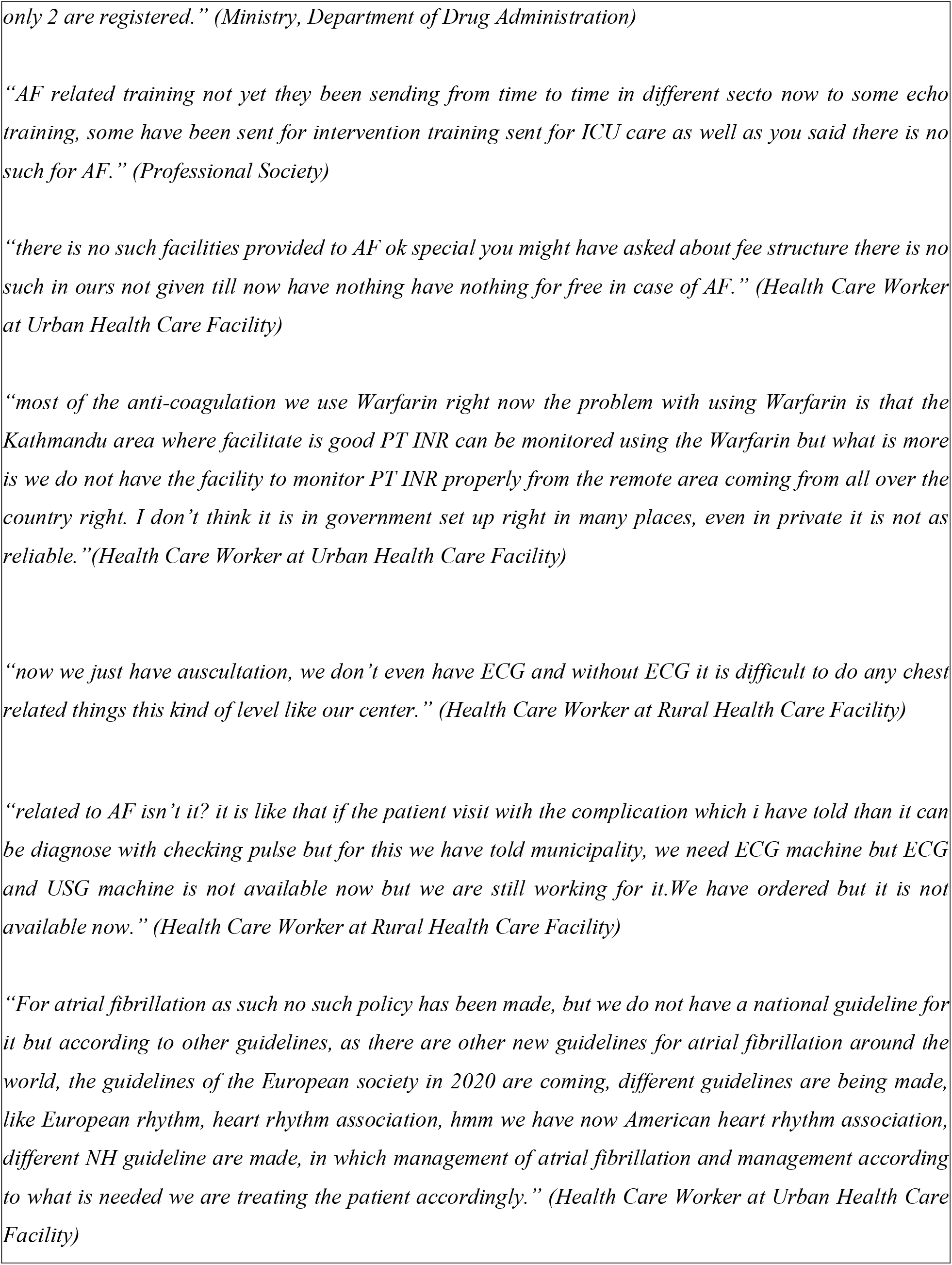
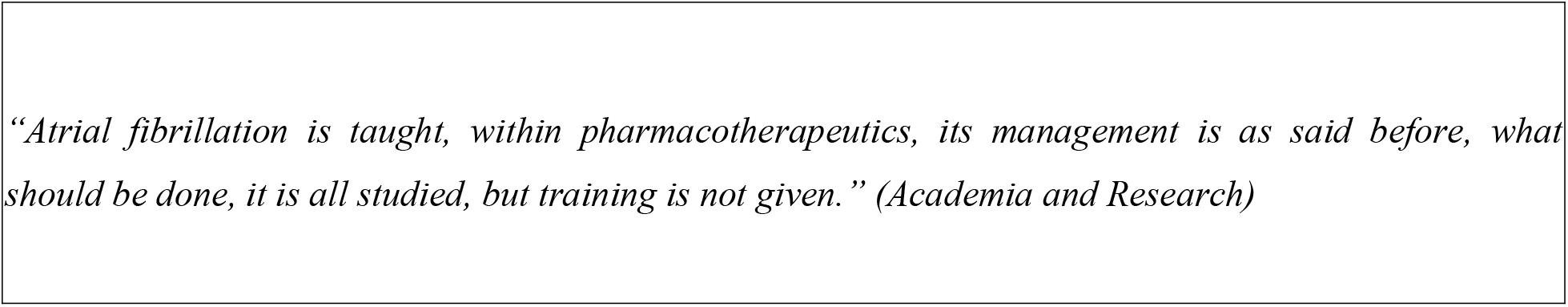
Quotes Describing Health Infrastructure, Guidelines and Training to Manage AF.

One of the participant (macro level) stated *“By making the protocol, hospitals up to PHC there is no need to send the protocol. Because there is no doctor as far as there is a doctor there is lab facility, till there if we can send atrial fibrillation standard protocol and now there are almost 200 hospitals and PHC in Nepal right. Whatever it is, let’s say it can be supplied by making a 300 chart, a flow chart of protocol. If that happens then atrial fibrillation will get the best result*.*” (Ministry)*

One of the other participant (macro level) emphasized the need of guideline by stating *“Talking about service from the perspective of drug and my observation of other places, while looking in the national level, there should be the standard treatment schedule protocol or guideline related to it. It should also include the procedure to diagnose and classification of drugs. We have multiple types of health facilities of different level in terms of resources and capacity. The guidelines should also specify the type/level of service to be provided from the different levels. And the first and foremost thing is that there should be a consistent guideline of national level*.*” (DDA)*

One of the participant (meso level) stated the difficulty in diagnosing and managing the case of AF due to inadequate trained healthcare professionals *“if you are telling manpower honestly we have health assistant like us for the diagnosis and treatment of AF. There must be staff nurse and medical officer, 3 satff will do that management, not really I don’t think AHW, ANM have been provided certain training and it is really difficult for them to diagnose and treat the patient with AF*.*” (Health Care Worker at Rural Health Care Facility)*

### Theme 3: Policy Environment for Cost Effective Management of AF with NOACs

There is no separate policy for AF management however there has been considerable work in designing and implementing policy for Cardiovascular Disease (CVD) risk and disease management by the government in the form of Package of Essential Non Communicable Diseases (PEN). PEN was introduced to screen, diagnose, treat and refer Cardio Vascular Diseases, chronic obstructive pulmonary disease, cancer, diabetes, and mental health at the grass root level of health system in for early detection and management of chronic diseases within the community. PEN however does not address management of AF directly but it does takes into consideration prevention and management of other CVD which can lead to AF.

The existing national policy includes 70 medicines into the essential list of drugs which are provided free of cost at the government hospital. The list includes medicines for basic management of hypertension and heart failure on the basis of programs like PEN package implemented in accordance with national health policy. There are medicines for management of arrhythmia but not VKAs, NOACS or antidote to NOACS taking into account cost of medicines for AF along the focus of national policy currently towards prevention of spectrum of CVDs. **(Table 5)**

**Table 5:**
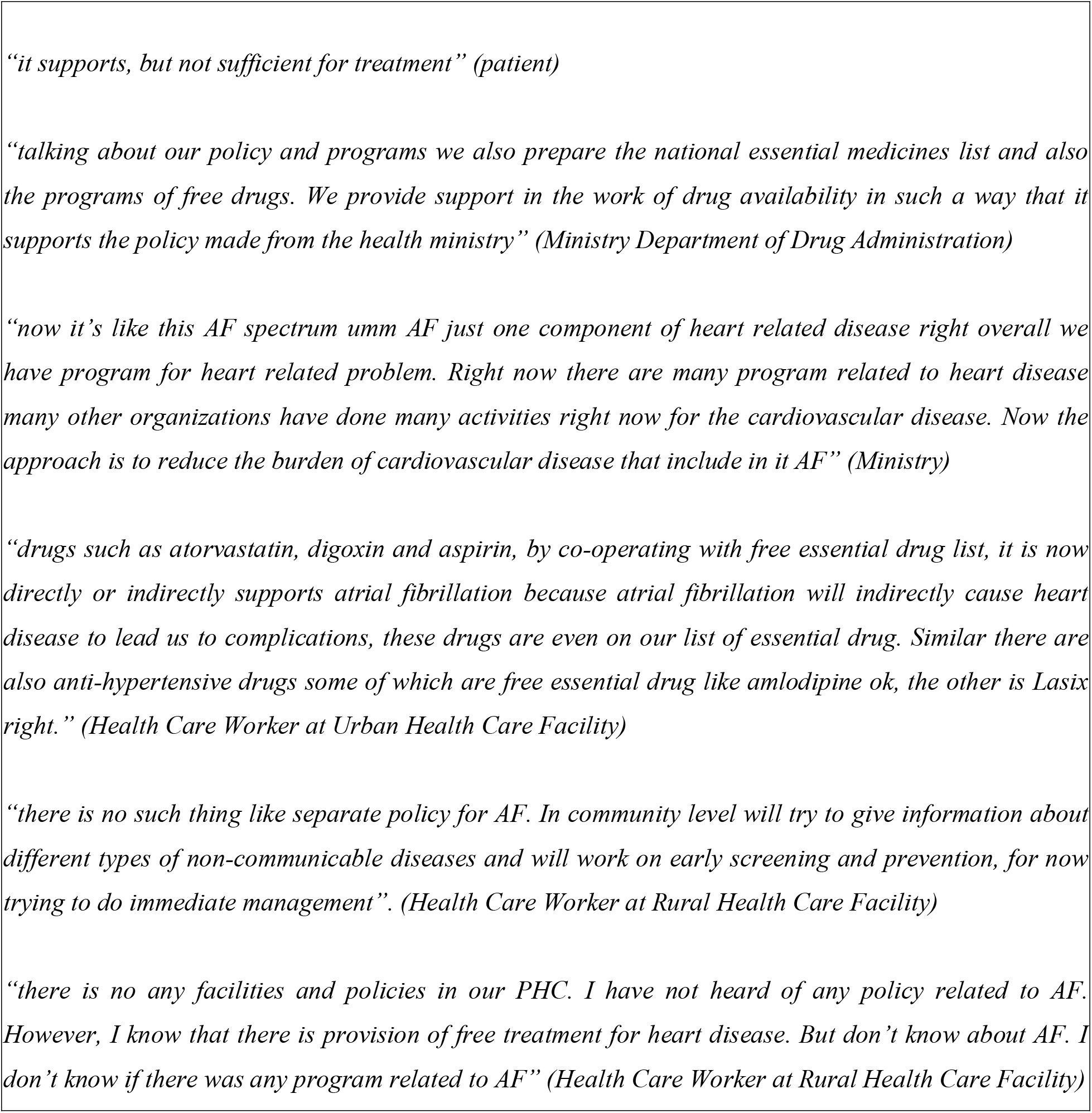
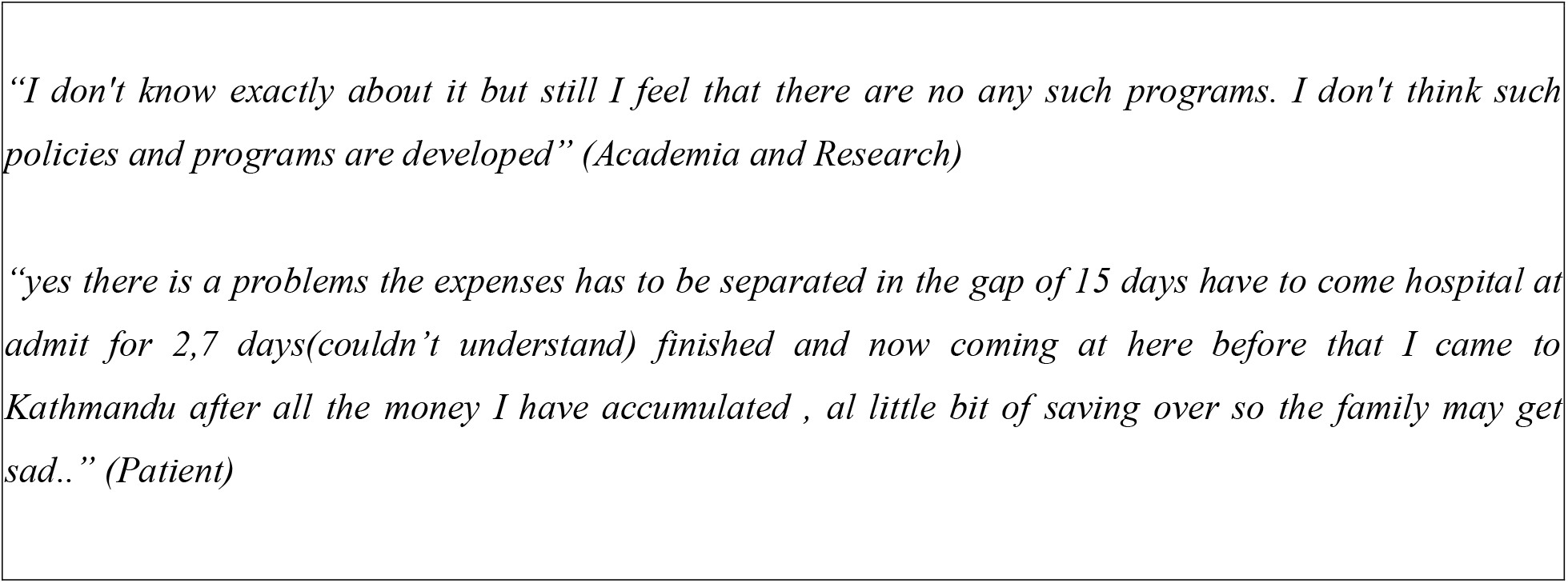
Policy Environment for Cost Effective Management of AF with NOACs.

**Table 6:**
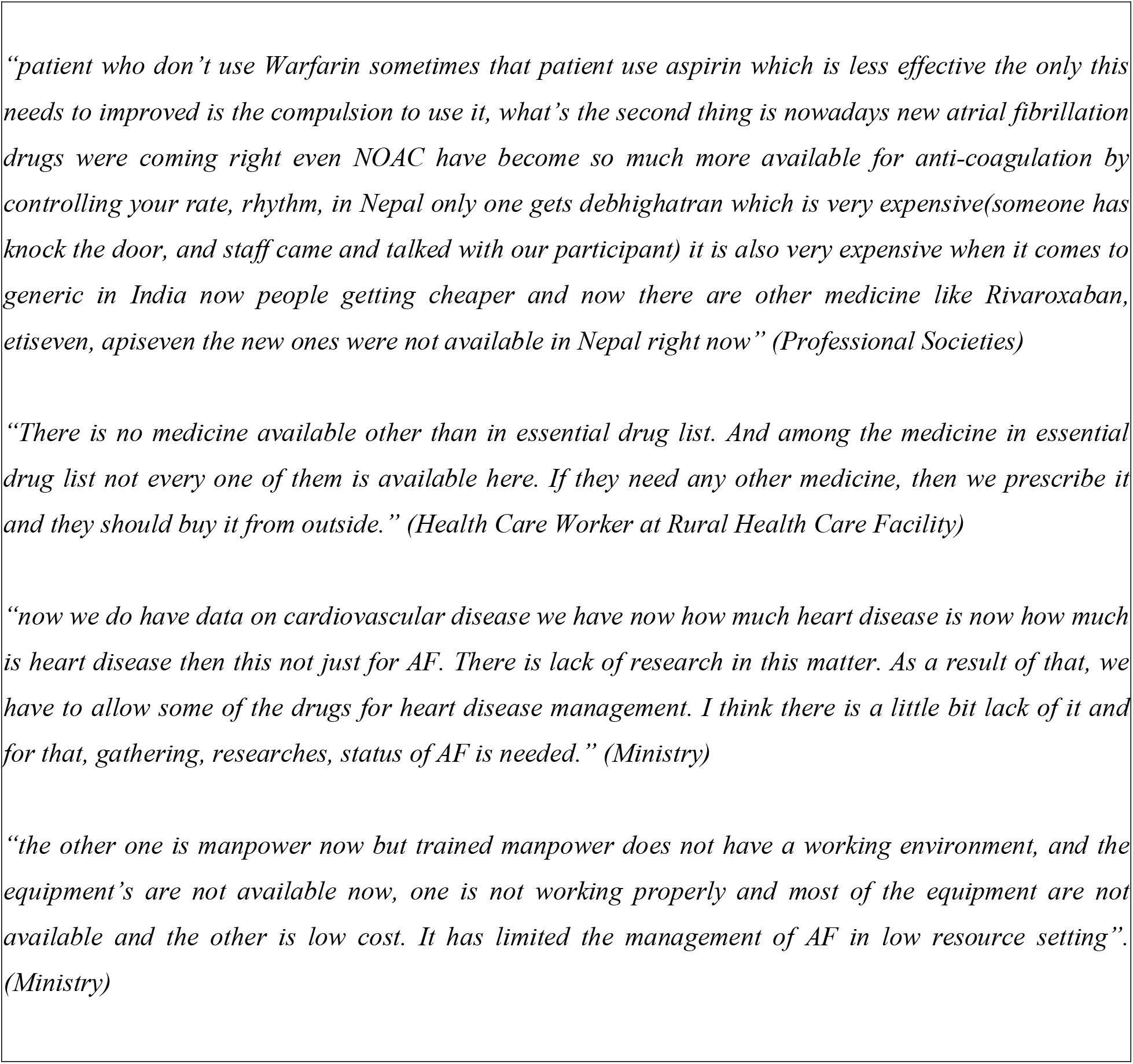
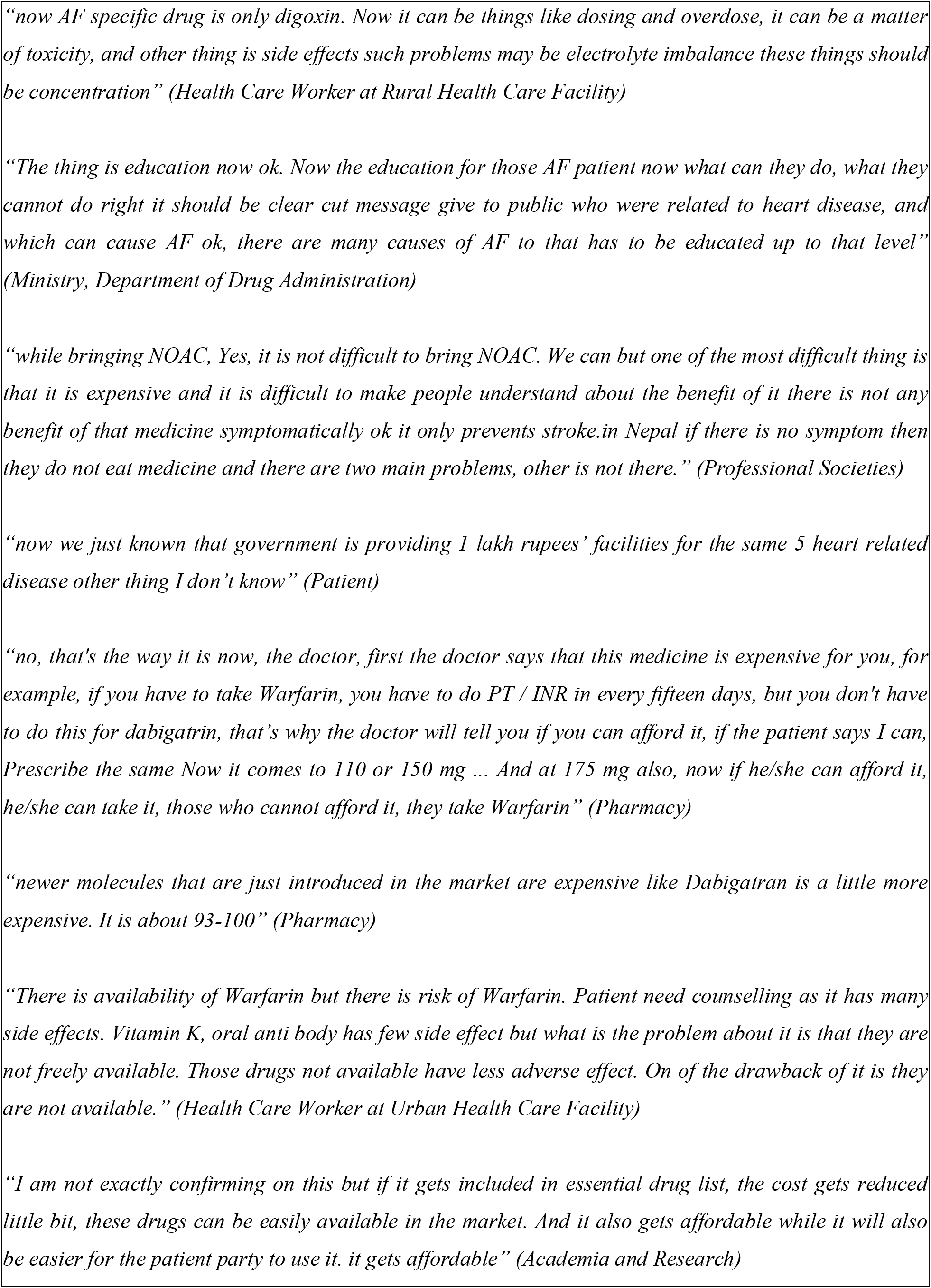
Barriers and Facilitators to Use of NOACs.

*“The essential drug list at present is in draft and this is the 6*^*th*^ *revision. Under cardiovascular we have amlodipine, in antihypertensive and at present we have not lept oral anti-coagulants in the anti-arrhythmic due to matter of cost. Till now we have not kept it due to cost factor. Till now there are Aarfarin, Aspirin, Clopidogrel, Furosemide. Likewise, we have diltiazem and diuretics, and we also have drugs of cancer*.*” (Academia and research)*. This statement form one of the micro level participants confirms that there is no inclusion of drugs to treat AF in the essential drug list which is available free of cost in government health care facilities.

One of the participant at macro level said *“we have a very high rate of mortality rate for this NCD. in this context, we have created a program on how to reduce the burden of NCD ok*… *Now WHO has now initiated the pen package as part of a program, the package of non-communicable disease is a training program that we are going to bring in the training program in 77 districts to build health workers’ knowledge till the end of this financial year and the training has running in 51 districts till now, we have separate them specially in 4 diseases and 4 risk factor. Among 4 diseases one is cardiovascular disease another one is diabetes and COPD and Cancer. (Ministry)* which indicates that the government in collaboration with WHO is working for CVD but not specific to AF

### Theme 4: Barriers and Facilitators to use of NOACS

The national essential drug list, which is provided free of cost at the government health care settings, includes drugs for the management of cardiovascular conditions like hypertension, MI, heart failure, stroke etc. however it does not include drugs for standalone treatment of AF. While VKAs are not available in the government and rural settings due to limited resources and infrastructure. Additionally, the rural settings are not equipped to regularly monitor INR, and the urban settings lack reliable monitoring centers.

Although the patients in urban areas might be willing to take NOACs owing to no requirement of INR monitoring, the availability of NOACs is limited in the urban healthcare settings and unavailable in the rural healthcare settings because of the high cost. Patients are not aware or counseled about routine INR monitoring when they are using VKAs and the patients switch to non VKAs like aspirin or leave the drug when they feel better which leads to further complications of AF. The majority of patients are unaware that they have AF but know they have a heart disease. Most of the patients were unaware of medications for heart conditions available free of cost in the rural government settings. These patients find the cost of regular medication and heart-related tests to be high, causing a significant financial burden for their households.

The national guideline focuses on managing the primary cardiovascular diseases with major national health programs and policies directed towards primary prevention. There is no guideline for management of AF alone and the drugs for AF albeit Digoxin which has many safety concerns are also not included in the essential drug list. One of the reason why national policy misses out on AF could be the lack of data on the national and subnational burden of AF.

One of the member of professional societies said *“mostly we use Warfarin right, now the problem with using Warfarin is that in Kathmandu (city) area where facilities is good PT INR can be monitored using the Warfarin but what is more is we do not have the facility to monitor PT INR properly from remote area coming from all over the country right. I do not think it is in government set up right in many places, even in private it is not as reliable*.*” (Professional Societies)*

*“I don’t think there are medicines for AF in primary, but we have to keep it in our essential list. But the ministry decides to which level it shall be kept. That’s because the AF diagnosis cannot be done in primary level. A specialist, trained health personnel is required to diagnose it, so the diagnosis cannot be done in primary*.*” (Ministry Department of Drug Administration)*

The patients experience financial problems with management of common heart conditions thus affording expensive medicine like NOAC adds on to that financial burden. *“there is a problem with expenses, have to come hospital and got admitted for 2,7 days and now all the money I have accumulated has finished, all saving is over so the family may get sad” (Patient)*

Making patient aware of AF, advantages and disadvantages of both VKAs and NOACs can facilitate better management of AF cases as stated by one of the medical practitioners in URBAN settings. *“But i think conducting the educational activities for follow up of AF, teaching the method for checking it on regular time interval, when to return to ER or OPD and correct use of VKAs and NOACs could help a little bit” (Health Care Worker at Urban Health Care Facility)*

*“For formulating policies and programs, in my opinion, firstly public awareness is very essential. While some of the people have asymptomatic AF some of them have paroxysmal AF. And what happens is that, there is chance that Asymptomatic AF becomes stroke later, but the patients don’t have any symptoms, so the patient party doesn’t want to go for checkup as they don’t feel any difficulties like that the other diseases make them feel. That’s why I feel public awareness is very much essential*.*” (Academia and Research)*

*“Likewise, in the present context, while doing various literature review, these oral anti coagulants are being underutilized. This novel anti-coagulant drug is expensive so it is not used so much. Though this Warfarin is given, it is being underutilized as its INR has to be maintained. The treatment is done from simply anti-platelet drugs such as aspirin due to the reason that it is difficult to monitor the INR as the patients from remote area cannot come frequently for the follow up. I feel that the health care facilities should be in the reach of many patient party so that they can have easy access to health. If only we can do that, the burden of the disease can be reduced. And the next thing is that I don’t think there are clinical guidelines, national guidelines of it*.*And if there will be also guidelines of it, the treatment would be proper. It would be even better if there will be policies related to it, if the policies will be developed in national level” (Academia and Research)*

## Discussion

NOACs are effective in preventing and treating thromboembolic conditions in patients with NVAF because they need less intense monitoring and have less drug-drug and drug-food interactions. Limited use of NOACs has been reported despite their proven effectiveness and the convenience they offer both to doctors for prescribing and to patients for usage. There are multitude of diverse factors via other priority programs (e.g. PEN), limited diagnostic infrastructure, no treatment guidelines, inadequate knowledge among health care workers, patients and care givers and various policy and health system barriers that affect the policy decision to include NOACS in the national EML. Patients may also not have knowledge of the disease of AF or its consequences if left untreated. Beyond the initial diagnosis and treatment, there are well-documented challenges with sustained adherence to recommended medications.

AF prevalence is underestimated in LMICs, like Nepal, mostly due to limited knowledge and awareness of AF and access to diagnostics. A recent study on physicians and nurse’s views towards the contributory factors related to NOACs medication errors identified lack of adequate knowledge and training, lack of confidence and lack of access to clinical guidelines and key factors related to NOACs errors.(18) Finding from this study also highlighted how physicians, mostly in the rural healthcare settings, lacked knowledge and training while physicians in both rural and urban settings lacked access to clinical national guidelines. As a result physicians continued the prescription of VKAs for treatment of NVAFs. Additionally, the patients lacked knowledge on symptoms of NVAFs and attributed the condition to some heart conditions.

One of the persistent challenges for the use of VKAs oral anticoagulants is the need of routine monitoring of PT/INR which has limitation from both health care providers and patients end. For healthcare providers, the primary limitation—across both urban and rural settings, as well as primary and secondary levels—is the lack of infrastructure and resources, including laboratories, reagents, and guidelines, necessary for assessing PT/INR.. Patients are generally unaware of NOACs and, at the same time, are unable to afford the cost of routine PT/INR monitoring. Given these circumstances, introducing NOACs into the national essential drug list seems plausible and important policy strategy for stroke prevention especially in the low middle income countries like Nepal. The findings of this study alligns with studies in various African countries which reported poor uptake of OACs due to reasons such as non-compliance of physicians to international recommendations on oral anticoagulation in patients with AF or limited accessibility and affordability of OACs, and the lack of resources for monitoring.(19,20)

Our findings indicate unavailability and suboptimal knowledge specific to AF training protocols while guidelines are not currently available for all HCPs (Health Care Providers) despite almost all health care workers felt there was a need for this. Additionally, lack of patient knowledge on the importance of continuous INR monitoring as a part of ideal management and on AF more generally were main barriers. The findings of qualitative meta-synthesis on HCPs views and experiences of prescribing anticoagulants for stroke prevention were consistent with the findings of our study which included HCPs lacking experience and confidence with prescribing and controlling anticoagulation. It also reported patients in HICS lacked information on the importance and impact of anticoagulation and often had misconceptions and misunderstandings about AF medications.(21)

The high cost of NAOCs is probably the most important barrier to the availability and widespread use of NOACS in a low and middle income countries like Nepal. A study in South Africa reported an estimated cost of 38 USD and 47 USD per month for Rivaroxaban and Dabigatran, respectively.(23). Despite the higher direct costs of NAOCs, they remain cost-effective for stroke prevention in patients with NVAF by reducing healthcare costs along with decreased incidence of stroke, major bleeding episodes, and associated consequences.(22) Some recommended interventions from the government stakeholders in this study were to lower the cost of essential medicines, giving patent to the national pharmaceuticals to ensure bulk purchasing with differential pricing for cost cutting. A cross-sectional survey conducted in India reported that the approximate monthly cost of Dabigatran at 150 mg twice a day to be □ 1500 (US$20) and Rivaroxaban at 20 mg once a day to be □ 3810 (US$52).(24) Furthermore, a cost-effectiveness analysis conducted in India showed that all direct costs of NAOCs, including Dabigatran and Rivaroxaban, lead to savings when compared with VKAs, resulting in a reduction of cost per patient.(25) In the UK, Dabigatran is priced at USD$65, while in China, it costs USD$222; similarly, Rivaroxaban is approximately USD$60 per patient per month in Kenya. (11) However, when considering expenses related to VKAs treatment, including low-priced medication (as low as USD$1 per month) and healthcare system costs like monitoring requirements, it indicates that over time, NOACs are c cost-effective.(11) However, the high variability in cost can limit access and causing NOACs unaffordable in certain low-income environments.

### Strength and Limitation of the study

This study incorporates diverse stakeholder perspectives to provide valuable insights into the facilitators and barriers to the use of NOAC. Employing a detailed qualitative approach, it explores barrier and facilitators through the perspectives of policymakers, healthcare providers, and patients, offering an understanding of NOAC adoption in Nepal’s resource-limited settings. As one of the few studies addressing this issue in a LMIC context, it identifies gaps in infrastructure, training, and awareness, providing actionable recommendations to improve AF management. However, the study has limitation too. As this study is limited to Bagmati Province, which may affect the generalizability of its findings to other regions with varying healthcare contexts. Additionally, despite the use of non-leading and non-judgmental questioning techniques, there is a possibility of social desirability bias in participant responses.

## Conclusion

The findings of the study shows that AF management in Nepal is hindered by limited awareness, inadequate infrastructure, lack of national guidelines, and restricted access to cost-effective medications. While health professionals in urban settings demonstrated better understanding of AF, rural practitioners and patients showed significant gaps in knowledge, particularly regarding the need for INR monitoring and the role of NOACs. The health system currently lacks standardized protocols and training dedicated to AF, and policy frameworks such as the PEN program address broader cardiovascular disease management but do not specifically target AF. Moreover, the absence of NOACs in the essential drug list, coupled with the high cost and poor accessibility, especially in rural areas, restricts optimal AF care. The findings of this study have significant implications for clinical practice and education, particularly among primary care physicians. There is an urgent toned to incorporate into the national educational/training program an overview of: the risks of atrial fibrillation and the anticoagulation under-treatment problem; accurate assessment of bleeding and stroke risk; benefits and disadvantages of anticoagulation options; prescription and insurance coverage guidelines for the available NOACs. The educational program could be embedded in a broader quality improvement initiative, in which all cadres of health care workers from the government health care setting should be included.

## Data Availability

All relevant data are within the manuscript and its Supporting Information files

## Acknowledgments

The authors would like to acknowledge all the participants of the study and all those who directly and indirectly helped us throughout our study.

